# Optimizing Lumipulse plasma pTau217/Aβ42 ratio cutpoints in an Asian cohort with high cerebrovascular disease burden

**DOI:** 10.64898/2026.11.04.26367313

**Authors:** Joyce R. Chong, Amelia T. Y. Yam, Anqi Toh, Gandan Jiang, Temmei Ito, Chizuru Kobayashi, Amitabh Dash, Pallavi Sachdev, Christopher P. Chen, Mitchell K. P. Lai

## Abstract

The generalizability of FDA-cleared Lumipulse plasma pTau217 / Aβ42 ratio cutpoints to Asian populations remains uncertain, especially in those with high burden of concomitant cerebrovascular disease (CeVD). We evaluated pTau217, Aβ42 and their ratio in 205 Singapore-based participants with corresponding amyloid neuroimaging. In-house cutpoints were derived using a three-range strategy targeting 95% sensitivity and specificity. Plasma pTau217/Aβ42 ratio outperformed pTau217 alone (area under the curve, 0.951 versus 0.925; p=0.004), with comparable performance across CeVD severities. Plasma ratio cutpoints achieved 98% negative predictive value, 88% positive predictive value, and 95% accuracy, classifying 10% as intermediate-risk. Using FDA-cleared cutpoints for our cohort retained 97% sensitivity and negative predictive value but yielded lower specificity (86%), positive predictive value (77%) and accuracy (85%), with 29% classified as intermediate-risk. Our findings support the need for population-specific validation of cutpoints from FDA-cleared blood biomarkers of brain amyloid pathology.

## 1. Introduction

The Lumipulse G pTau217/Aβ42 plasma ratio from Fujirebio (Tokyo, Japan) is the first US Food and Drug Administration (FDA)-cleared blood-based marker of amyloid pathology associated with Alzheimer’s disease in symptomatic older adults [1]. Although the FDA-reviewed multi-center study reported good diagnostic performance of the published cutpoints [2], an important caveat remains: the cohort was predominantly White (85%), and the generalizability of these cutpoints to non-White populations remains under-studied, particularly amongst individuals with differing sociodemographic characteristics and comorbidities that may influence plasma biomarker profiles [3-5]. Here, using a well characterized, longitudinally assessed Asian cohort with high concomitant cerebrovascular disease (CeVD) burden and vascular risk factors [6], we evaluated Lumipulse pTau217/Aβ42 plasma ratio and pTau217 alone for detection of amyloid positivity (Aβ+) on positron emission tomography (PET), derived in-house cutpoints using a three-range strategy, and compared their performance with FDA-cleared cutpoints.

## 2. Methods

### 2.1. Study population

Participants in the current study were a subset of a longitudinal cohort from the National University Hospital Memory Clinic and the community in Singapore [6] for whom brain amyloid status had been measured. Clinical diagnoses included cognitively normal (CN), cognitively impaired no dementia (CIND, defined as participants with impairment in at least one cognitive domain upon formal neuropsychological assessments, but who did not meet the Diagnostic and Statistical Manual of Mental Disorders-Fourth Edition (DSM-IV) criteria for dementia), Alzheimer’s disease (AD, fulfilling DSM-IV criteria for dementia as well as the National Institute of Neurological and Communicative Disorders and Stroke and the Alzheimer’s Disease and Related Disorders Association [NINCDS-ADRDA] criteria [7]), or vascular dementia (VaD, fulfilling DSM-IV criteria for dementia as well as the National Institute of Neurological Disorders and Stroke and Association Internationale pour la Recherché et l’ Enseignement en Neurosciences [NINDS-AIREN] criteria [8]). Additionally, demographic data and comprehensive measures of cognitive function, AD and vascular risk factors as well as CeVD have been previous described [9, 10] and are detailed in **Supplementary Table 1**.

### 2.2. Amyloid PET quantification

Determination of brain amyloid positivity (Aβ+) by visual assessments of amyloid PET has been previously described [9, 10]. Briefly, Images scanned with [^11^C]-Pittsburgh Compound B (PiB) or [^18^F]-flutafuranol on an mMR synchronous PET/MR scanner (Siemens Healthcare GmbH, Munich, Germany) were reconstructed using ordinary Poisson ordered-subsets expectation maximization with all corrections applied, with each image visually interpreted independently as Aβ+ or Aβ-by three experienced raters blinded to the clinical diagnosis of participants.

### 2.3. Blood collection and assays

Blood from non-fasted participants was collected into ethylenediaminetetraacetic acid (EDTA) tubes and centrifuged at 2000 *g* for 10 minutes at 4°C. Plasma was extracted, aliquoted and stored at −80°C until use. The chemiluminescent enzyme immunoassay-based Lumipulse® G assays for plasma pTau217 (tau phosphorylated at threonine 217) and Aβ42 (recognizing β-amyloid_1-42_ peptide) were performed by Fujirebio Diagnostics Inc.’s subsidiary in Tokyo, Japan (reagent lots: D4C6086J and C5C6091S) and expressed in units of pg/mL. Evaluators were blinded to clinical information.

### 2.4. Statistical analysis

Differences in plasma biomarkers among groups stratified by clinical diagnosis and Aβ-PET status were assessed using Kruskal-Wallis analyses of variance and *post-hoc* Dunn’s tests. Diagnostic performance for detecting PET Aβ+ was assessed using area under the receiver-operating characteristic curve (AUC). P-values for comparison of AUCs were derived from DeLong tests. In-house pTau217/Aβ42 and pTau217 cutpoints for PET Aβ+ were derived using a three-range strategy comprising a lower reference point to rule out AD (95% sensitivity; low-risk for PET Aβ+) and a higher reference point to rule in AD (95% specificity; high-risk for PET Aβ+). Participants with values between the reference points were classified as having intermediate-risk. All in-house and FDA-cleared cutpoints are presented in **Table 1 and Figure 1A, 1B**.

**Fig. 1.**
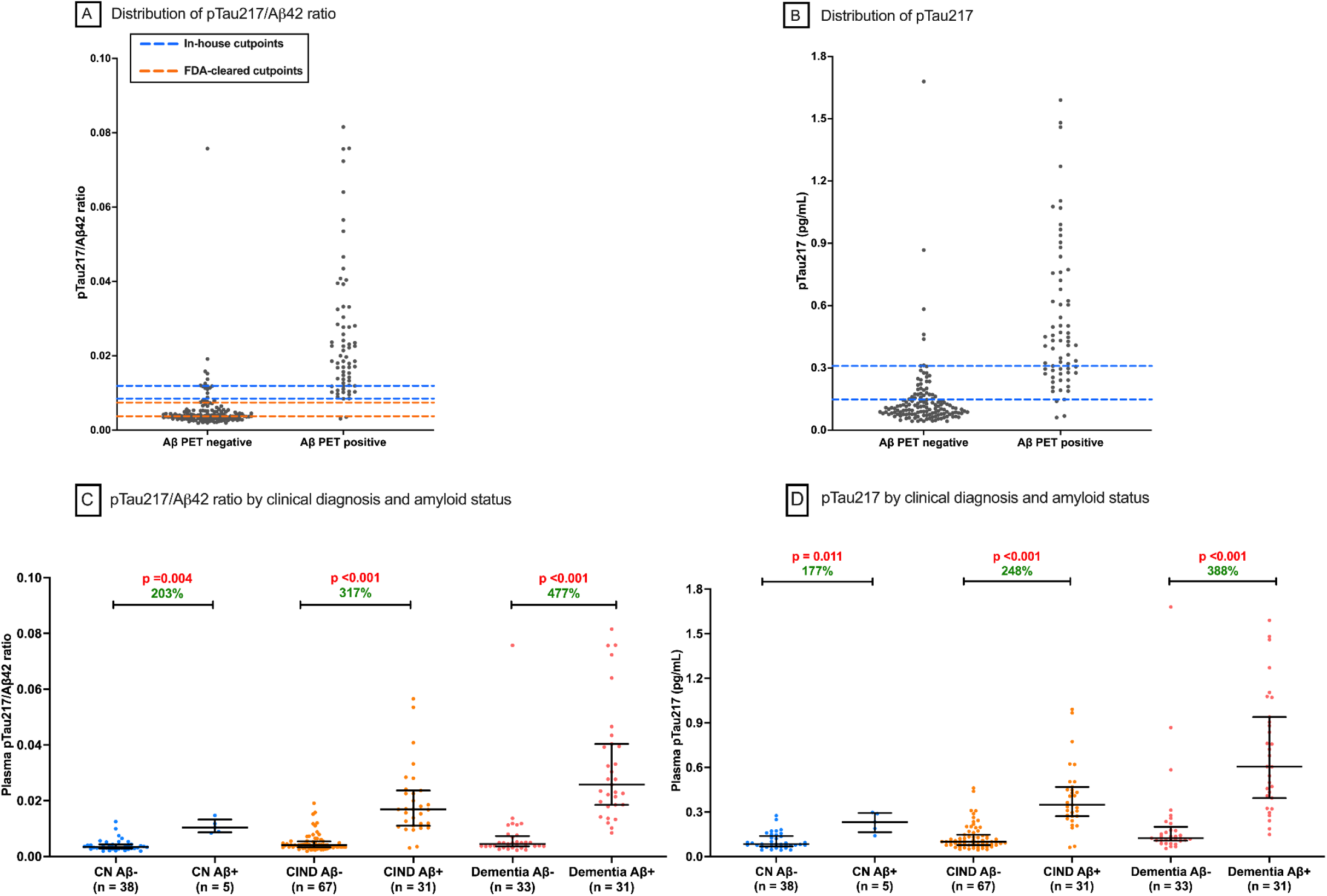
Lumipulse pTau217/Aβ42 Ratio and pTau217 by Clinical Diagnosis and Amyloid Status. *Abbreviations*: CN, cognitively normal; CIND, cognitive impairment no dementia; FDA, US Food and Drug Administration; PET, positron emission tomography. **A**. Plot depicts distribution of plasma pTau217/Aβ42 ratio in PET amyloid-negative and amyloid-positive individuals. For pTau217/Aβ42 ratio, the in-house cutpoints for risk of Aβ-PET positivity are as follows: 0.00846 or lower indicates low-risk; 0.00847 to 0.01188, intermediate-risk; and 0.01189 or higher, high-risk. The FDA-cleared cut points are as follows: 0.00370 or less, negative; 0.00371 to 0.00737, indeterminate; and 0.00738 or higher, positive. **B**. Plot depicts distribution of plasma pTau217 concentration in PET amyloid-negative and amyloid-positive individuals. For pTau217 concentration (pg/mL), the in-house cutpoints for risk of Aβ-PET positivity are as follows: 0.1481 or lower indicates low-risk; 0.1482 to 0.3093, intermediate-risk; and 0.3094 or higher, high-risk. **C and D**. Plots depict distributions of plasma pTau217/Aβ42 ratio and pTau217, respectively, by clinical diagnosis and Aβ-PET status. The graphs show the median and interquartile range. p-values derived from Kruskal–Wallis tests followed by post hoc Dunn’s tests. Within each clinical diagnostic group, the percentage increase of plasma biomarker levels in the amyloid-positive (Aβ+) group from the amyloid-negative (Aβ–) group is shown in green (percentage increase = [(Median Aβ+ - Median Aβ-) / Median Aβ-] × 100%). Red indicates significant p-values.

**Table 1.** Performance Characteristics of the Lumipulse pTau217/Aβ42 Ratio and pTau217.

|  | pTau217/Aβ42 |  | pTau217 |
|---|---|---|---|
|  | **In-house cutpoints** <sup>a</sup><br>for Aβ-PET positivity:<br>**pTau217/Aβ42 ≥ 0.01189,<br>pTau217/Aβ42 ≤ 0.00846** | **FDA-cleared cutpoints** <sup>b</sup><br>for Aβ-PET positivity:<br>**pTau217/Aβ42 ≥ 0.00738,<br>pTau217/Aβ42 ≤ 0.00370** | **In-house cutpoints** <sup>a</sup><br>for Aβ-PET positivity:<br>**pTau217 ≥ 0.3094,<br>pTau217 ≤ 0.1481** |
| **Classification** |  |  |  |
| **Low-risk, n (%)** | 126 (61) | 61 (30) | 105 (51) |
| PET Aβ-, n | 123 | 59 | 102 |
| PET Aβ+, n | 3 | 2 | 3 |
| **Intermediate-risk, n (%)** | 20 (10) | 59 (29) | 48 (24) |
| PET Aβ-, n | 8 | 59 | 29 |
| PET Aβ+, n | 12 | 0 | 19 |
| **High-risk, n (%)** | 59 (29) | 85 (41) | 52 (25) |
| PET Aβ-, n | 7 | 20 | 7 |
| PET Aβ+, n | 52 | 65 | 45 |
| **Sensitivity, %** | 95.5 | 97.0 | 95.5 |
| **Specificity, %** | 94.9 | 85.5 | 94.9 |
| **NPV for Low-risk, %** <sup>c</sup> | 97.6 | 96.7 | 97.1 |
| **PPV for High-risk, %** <sup>c</sup> | 88.1 | 76.5 | 86.5 |
| **Accuracy for Low- and High-risk, %** <sup>c</sup> | 94.6 | 84.9 | 93.6 |

*Abbreviations*: FDA, US Food and Drug Administration; NPV, negative predictive value; PET, positron emission tomography; PPV, positive predictive value.
<sup>a</sup> The in-house cutpoints for Aβ-PET positivity were derived based on 95% sensitivity (lower reference point) and 95% specificity (upper reference point).
<sup>b</sup> The FDA-cleared Lumipulse pTau217/Aβ42 ratio categorizes results as negative, indeterminate, or positive, corresponding respectively to low-risk, intermediate-risk, and high-risk classifications.
<sup>**c**</sup> Participants with intermediate results were excluded from diagnostic performance calculations of NPV, PPV, and accuracy.

## 3. Results

205 participants were included (mean [SD] age, 75.7 [6.9] years; 118 [58%] female; 43 cognitively normal [CN], 98 cognitively impaired no dementia [CIND], 64 dementia; 67 were PET Aβ+ [33%]; 145 had significant CeVD [73%]; see **Supplementary Table 1**). Both pTau217/Aβ42 and pTau217 were significantly higher in Aβ+ participants compared with Aβ-participants, across all diagnostic groups (all p’s ≤ 0.011; **Figure 1C and 1D**). Compared with pTau217, pTau217/Aβ42 showed higher percentage increase between Aβ- and Aβ+ participants in all clinical stages.

Plasma pTau217/Aβ42 (AUC [95% CI] = 0.951 [0.917-0.985]) outperformed pTau217 alone (AUC [95% CI] = 0.925 [0.881-0.968]) in detecting PET Aβ+ (p = 0.004). In sensitivity analyses, among dementia-free participants (CN + CIND), pTau217/Aβ42 also achieved a high AUC of 0.936 (95% CI, 0.883-0.990), outperforming pTau217 (AUC [95% CI] = 0.911 [0.844-0.978]; p = 0.035). Next, pTau217/Aβ42 performed comparably in CeVD- and CeVD+ individuals, demonstrating good performance in both groups (AUC [95% CI]; CeVD-: 0.967 [0.906-1.00]; CeVD+: 0.940 [0.898-0.983]; p = 0.48).

Using in-house pTau217/Aβ42 cutpoints (95% sensitivity, 95% specificity), low- and high-risk classifications achieved negative predictive value (NPV) of 98%, positive predictive value (PPV) of 88%, and accuracy of 95% (**Table 1**). The proportion of participants in intermediate-risk group was 10%. Using FDA-cleared pTau217/Aβ42 cutpoints, sensitivity and NPV remained high at 97%. However, specificity decreased to 86%, PPV to 77%, and accuracy to 85%. Additionally, the proportion with intermediate- or indeterminate risk was substantially higher at 29%. In-house pTau217 cutpoints yielded NPV, PPV, and accuracy of 97%, 87%, and 94%, respectively, with 24% of participants in intermediate-risk group.

To identify potential factors associated with discrepancies between classifications based on FDA-cleared and in-house cutpoints, we compared demographic and comorbidity profiles of participants with concordant versus discordant risk group classifications. The discordant group had a lower proportion of females (38% vs 72%; p < 0.001 by chi-square test) and higher proportions of participants with diabetes (35% vs 23%; p = 0.048) and chronic kidney disease (76% vs 61%; p = 0.025).

## 4. Discussion

With the advent of ultrasensitive platforms, the field of dementia blood-based biomarkers research is advancing rapidly, currently reaching a consensus that plasma phosphorylated tau species are associated with brain amyloid burden, with pTau217 generally demonstrating higher accuracy for detecting amyloid pathology compared to other pTau species [11, 12]. This consensus is reflected by three out of four of the currently FDA-cleared blood tests incorporating pTau217, either by itself (Elecys pTau217 from Roche), or combined with Aβ (PrecivityAD2 from C2N Diagnostic as well as Lumipulse). As the first FDA-cleared blood test, the Lumipulse plasma pTau217/Aβ42 ratio has had some lead time for characterization, but the majority of studies remain focused on western populations. Here, we have demonstrated excellent performance of the Lumipulse plasma pTau217/Aβ42 ratio in detecting abnormal brain amyloid burden in an Asian cohort with high prevalence of concomitant CeVD (AUC = 0.951). While both in-house and FDA-cleared pTau217/Aβ42 cutpoints achieved high NPV and sensitivity of ≥ 95%, the in-house cutpoints outperformed FDA-cleared cutpoints by reducing the proportion of intermediate-risk results (29% vs. 10%) and improving PPV, specificity, and accuracy. Importantly, a classification of intermediate-risk necessitates confirmatory tests which are more costly and invasive (e.g., CSF tests and amyloid PET), and the Global CEO Initiative on Alzheimer’s Disease workgroup has recommended that for a blood-based biomarker test to be useful in the clinical setting for amyloid pathology risk-stratification, the proportion of intermediate-risk in three-range cutoffs should be no more than 15-20% in a typical clinical population [13]. In our cohort, using the in-house cutpoints met the recommended threshold, whilst using FDA-cleared cutpoints did not.

There is further evidence of cohort differences from the few other non-White studies published for the Lumipulse assay. For example, similar to our findings, a Japanese study reported decreases of intermediate-risk proportions when locally optimized thresholds were used instead of the FDA-cleared cutpoints [14]. In contrast, a head-to-head comparison of Black / African American vs. White participants showed high and comparable performance across the two races [15]. Taken together, these findings underscore the importance of validating biomarker performance and cutpoints, as well as comparisons with FDA-cleared cutpoints, in cohorts that resemble the intended-use populations as blood biomarkers enter routine clinical practice. Furthermore, as newer biomarkers (e.g., brain-derived pTau and eMTBR-tau243 [10, 16, 17]) enter evaluation phases, it may be worthwhile to diversify the cohorts under validation both ethnically and geographically.

Finally, given the single-cohort design of this study, further validation is warranted in other Asian cohorts and under-represented populations, where differences in demographic characteristics and comorbidity profiles, or preanalytical and analytical factors [18], may influence the performance and applicability of blood biomarkers and their cutpoints [5].

## Data Availability

When published in final form, all data will be available from the corresponding author upon reasonable request.

## Ethics approval and consent to participate

Study approval was obtained from Singapore National Healthcare Group Domain-Specific Review Board (2018/00996, 2015/00406, and 2015/00441). Written informed consent was obtained from study participants or their next-of-kin.

## Consent for publication

Not applicable. This manuscript does not contain any individual person’s data in any form.

## Declaration of the use of generative AI and AI-assisted technologies

We have nothing to disclose.

## CRediT authorship contribution statement

**Joyce Chong:** Conceptualization, Methodology, Formal analysis, Validation, Data curation, Writing – Original Draft. **Amelia Yam:** Formal analysis, Project administration, Writing – Review & Editing. **Anqi Toh:** Data curation, Writing – Review & Editing. **Gandan Jiang:** Investigation, Resources, Writing – Review & Editing. **Temmei Ito:** Investigation, Resources, Writing – Review & Editing. **Chizuru Kobayashi:** Investigation, Resources, Writing – Review & Editing. **Amitabh Dash:** Investigation, Resources, Writing – Review & Editing. **Pallavi Sachdev:** Investigation, Resources, Writing – Review & Editing. **Christopher Chen:** Conceptualization, Methodology, Supervision, Funding acquisition, Writing – Review & Editing. **Mitchell Lai:** Conceptualization, Validation, Data curation, Supervision, Funding acquisition, Writing – Original Draft.

## Acknowledgments

We would like to thank the study patients and their caregivers for their support.

## Supplementary materials

Supplementary Table 1. Characteristics of study participants: can be found linked to this article.

## References

1. US Food and Drug Administration. 510(k) Premarket Notification: Immunoassay blood test for amyloid pathology assessment. June 30, 2026]; Available from: https://www.accessdata.fda.gov/scripts/cdrh/cfdocs/cfpmn/pmn.cfm?ID=K242706.

2. Moghekar, A.R., et al., Multicenter validation of plasma p-tau217/ amyloid beta 1-42 ratio in symptomatic Alzheimer’s disease. Alzheimers Dement, 2026. 22(7): p. e71644.

3. Chong, J.R., et al., Association of plasma GFAP with elevated brain amyloid is dependent on severity of white matter lesions in an Asian cognitively impaired cohort. Alzheimers Dement (Amst), 2024. 16(2): p. e12576.

4. Lam, B.Y.K., et al., High burden of cerebral white matter lesion in 9 Asian cities. Sci Rep, 2021. 11(1): p. 11587.

5. Schöll, M., et al., Challenges in the practical implementation of blood biomarkers for Alzheimer’s disease. Lancet Healthy Longev, 2024. 5(10): p. 100630.

6. Lim, M.J., et al., HARMONISATION - A multimodal prospective study of vascular cognitive impairment in multi-ethnic Asians: Cohort profile, progress, current contributions, and future impact. J Alzheimers Dis, 2025. 108(4): p. 1452–1474.

7. McKhann, G., et al., Clinical diagnosis of Alzheimer’s disease: report of the NINCDS-ADRDA Work Group under the auspices of Department of Health and Human Services Task Force on Alzheimer’s Disease. Neurology, 1984. 34(7): p. 939–44.

8. Román, G.C., et al., Vascular dementia: diagnostic criteria for research studies: report of the NINDS□AIREN International Workshop. Neurology, 1993. 43(2): p. 250–250.

9. Chong, J.R., et al., Clinical utility of plasma p-tau217 in identifying abnormal brain amyloid burden in an Asian cohort with high prevalence of concomitant cerebrovascular disease. Alzheimers Dement, 2025. 21(2): p. e14502.

10. Chong, J.R., et al., Plasma brain-derived p-Tau217 outperforms other p-Tau species in detecting abnormal brain amyloid in an Asian cohort of older people with cerebrovascular disease burden. J Prev Alzheimers Dis, 2026. 13(7): p. 100615.

11. Mendes, A.J., et al., Head-to-head study of diagnostic accuracy of plasma and cerebrospinal fluid p-tau217 versus p-tau181 and p-tau231 in a memory clinic cohort. J Neurol, 2024. 271(4): p. 2053–2066.

12. Janelidze, S., et al., Head-to-head comparison of 10 plasma phospho-tau assays in prodromal Alzheimer’s disease. Brain, 2023. 146(4): p. 1592–1601.

13. Schindler, S.E., et al., Acceptable performance of blood biomarker tests of amyloid pathology - recommendations from the Global CEO Initiative on Alzheimer’s Disease. Nat Rev Neurol, 2024. 20(7): p. 426–439.

14. Ishiguro, T., et al., Prospective study on clinical utility of plasma p-Tau217 and other biomarkers in Japanese memory clinics using the LUMIPULSE platform. Alzheimers Res Ther, 2026. 18(1).

15. Cousins, K.A.Q., et al., Comparison of plasma p-tau217/Aβ42, p-tau217, and Aβ42/Aβ40 biomarkers by race to detect Alzheimer’s disease. Alzheimers Dement, 2025. 21(8): p. e70469.

16. Horie, K., et al., Plasma MTBR-tau243 biomarker identifies tau tangle pathology in Alzheimer’s disease. Nat Med, 2025. 31(6): p. 2044–2053.

17. Denkinger, M.N., et al., Brain-derived plasma p-tau217 shows enhanced dynamic range for Alzheimer’s disease neuropathological change. Alzheimers Dement, 2026. 22(9): p. e71773.

18. Algeciras-Schimnich, A., et al., Variability in Clinical Performance of the FDA-Cleared Lumipulse P-Tau217/β-Amyloid 1–42 Plasma Ratio. JAMA Neurology, 2026.

