## Supplementary Table 1 for "Optimizing Lumipulse plasma pTau217/Aβ42 ratio cutpoints in an Asian cohort with high cerebrovascular disease burden"

**Supplementary Table 1. Characteristics of study participants**

| | Aβ PET negative (Aβ–) [a] | Aβ PET positive (Aβ+) [a] | p-value [b] | n available |
|---|---|---|---|---|
| n | 138 (67) | 67 (33) | | |
| Age, years, mean (SD) | 75.0 (6.8) | 77.2 (6.9)* | 0.033 | 205 |
| Female, n (%) | 67 (49) | 51 (76)* | <0.001 | 205 |
| Education, years, mean (SD) | 8.1 (4.9) | 7.3 (5.0) | 0.272 | 205 |
| APOE ε4 carrier, n (%) | 26 (19) | 32 (48)* | <0.001 | 205 |
| MMSE, median (IQR) | 25 (6) | 19 (9)* | <0.001 | 205 |
| **Clinical diagnoses [c]** | | | <0.001 | 205 |
| CN, n (%) | 38 (27) | 5 (8)* | | |
| CIND, n (%) | 67 (49) | 31 (46) | | |
| AD, n (%) | 15 (11) | 28 (42)* | | |
| VaD, n (%) | 18 (13) | 3 (4) | | |
| **Comorbidity** | | | | |
| Hypertension, n (%) | 103 (75) | 49 (73) | 0.818 | 205 |
| Hyperlipidemia, n (%) | 103 (75) | 47 (70) | 0.496 | 205 |
| Diabetes, n (%) | 41 (30) | 16 (24) | 0.416 | 204 |
| Cardiovascular disease, n (%) | 12 (10) | 4 (6) | 0.408 | 189 |
| Chronic kidney disease, n (%) | 93 (69) | 40 (63) | 0.333 | 198 |
| BMI, n (%) | | | 0.421 | 205 |
| Underweight (≤18.4) | 10 (7) | 5 (8) | | |
| Normal (18.5 to 27.4) | 99 (72) | 53 (79) | | |
| Obese (≥27.5) | 29 (21) | 9 (13) | | |
| **Significant CeVD, n (%) [d]** | 96 (73) | 49 (74) | 0.820 | 198 |
| **Plasma biomarkers** | | | | |
| pTau217, pg/mL, median (IQR) | 0.103 (0.08) | 0.428 (0.4)* | <0.001 | 205 |
| pTau217/Aβ42 ratio, median (IQR) | 0.00385 (0.002) | 0.0185 (0.02)* | <0.001 | 205 |

*Abbreviations:* AD, Alzheimer's disease; BMI, body mass index; CeVD, cerebrovascular disease; CIND, cognitive impairment no dementia; CN, cognitively normal; IQR, interquartile range; MMSE, Mini-Mental State Examination; PET, positron emission tomography; SD, standard deviation; VaD, vascular dementia.

[a] Amyloid PET status was based on visual read of [$^{11}$C]PiB (n = 175) or [$^{18}$F]Flutafuranol (n = 30) imaging.
[b] p-values were derived from Chi-square tests for categorial variables, and from independent-sample t-test or Mann–Whitney U test for normally distributed or skewed continuous variables, respectively.

[c] Participants were classified at research consensus meetings with study clinicians and psychologists into one of the following diagnostic categories: (1) cognitively normal (CN): individuals without objective cognitive impairment on formal neuropsychological assessments, or functional loss; (2) cognitive impairment no dementia (CIND): individuals with impairment in at least one cognitive domain upon formal neuropsychological assessments, but who did not meet the Diagnostic and Statistical Manual of Mental Disorders, Fourth Edition (DSM-IV) criteria for dementia; (3) dementia: individuals diagnosed according to the DSM-IV criteria, with further etiologic diagnoses following the National Institute of Neurological and Communicative Disorders and Stroke and the Alzheimer's Disease and Related Disorders Association (NINCDS-ADRDA) criteria for Alzheimer's disease (AD), and the National Institute of Neurological Disorders and Stroke and Association Internationale pour la Recherché et l' Enseignement en Neurosciences (NINDS-AIREN) criteria for vascular dementia (VaD).
[d] Significant CeVD was defined as total number of cortical infarcts ≥1, and/or total number of lacunes ≥2 and/or total number of cerebral microbleeds ≥2 and/or Fazekas score ≥2.

* Significant difference between Aβ PET negative and Aβ PET positive.
